# Instantaneous R calculation for COVID-19 epidemic in Brazil

**DOI:** 10.1101/2020.04.23.20077172

**Authors:** Francisco H. C. Felix, Juvenia Fontenele

## Abstract

COVID-19 pandemic represents a major challenge to health systems of all countries. Brazilian regions habe been showing marked differences in onset and number of cases. Health authorities instituted widespread social distancing and lockdown measures but their implementation has also varied. The authors used data on confirmed cases of COVID-19 in Brazil and its states to calculate the value of instantaneous reproduction number at these regions. The results show a reduction of instantaneous reproduction number with time, probably due to social distancing measures put in place in the last weeks by brazillian authorities. It seems logical to maintain restrictions to social contact until the epidemic peak has occurred in Brazil.

## Introduction

On December 31, 2019, 27 cases of viral pneumonia were reported in the city of Wuhan, China. A new coronavirus, related to SARS-Cov and MERS-Cov, was isolated from the patients’ airways, being initially named 2019-nCov (Zhu et al., 2020). The rapid spread of this new coronavirus (now called SARS-Cov-2) gave rise to the current pandemic, (Bedford et al., 2020). If there is a significant number of asymptomatic patients transmitting the virus isolation of only the high-risk population will be ineffective (Hellewell et al., 2020). Widespread social restrictions, therefore, are necessary for the effective control of the pandemic (Anastas-sopoulou et al., 2020; Hou et al., 2020).

Outbreaks can be quantitativelly described by the reproduction number (R) (Heesterbeek and Dietz, 1996; Delamater et al., 2019) a common measure of pathogen spread. R values higher than 1 are associated with increasing spread and epidemic phase, whereas decline of spread is seen with values of less than 1. An estimate of COVID-19 R_0_ value (basic reproduction number on the initial outbreak) was made using Wuhan data, calculating a number between 2.2 and 3.6 (Li et al., 2020). The first brazillian case was registered on February 26, 2020. As of today (April 16), the total number of confirmed cases is 30,425. It is still unclear what the epidemic dynamics are locally and how it has responded to social restrictions.

## Methods

### Data source

The data were obtained from official sources. The number of cumulative confirmed total cases and deaths in Ceará was obtained from the Ceará State Government information site, IntegraSUS (do Estado do Ceará, 2020). The number of cumulative confirmed total cases and deaths in the state of São Paulo was obtained from the coronavirus information site in the state of São Paulo (do Estado de São Paulo, 2020). The number of cumulative confirmed total cases and deaths in Brazil and its other states was obtained from the COVID-19 brazillian information website (da Saúde do Brasil, 2020). We used the number of cumulative active cases (total minus deaths) to build the epidemic curves.

### Statistical analysis

We used EpiEstim, a web application to estimate disease transmissibility during an infectious disease outbreak from incidence time series, computing time-dependent reproduction number (R) (Thompson et al., 2019). The Wallinga and Teunis method, as implemented by Ferguson (Wallinga, 2004; Cori et al., 2013) is a likelihood based estimation procedure that captures the temporal pattern of effective reproduction numbers from an observed epidemic curve.

Instantaneous R (*R*_*i*_) was estimated within a 5 day time window. Prior mean and standard deviation values for R were set at 3 and 1. Serial interval was estimated using a parametric distribution with uncertainty (offset gamma). We compared the results at two time points (day 7 and day 21 after the first case was registered at each region) from different brazillian states in order to make inferences about the epidemic dynamics.

## Results

The evaluation of *R*_*i*_ at the first time point (7 days after the first case was registered in each state) showed a variation in the range of 1.55 to 2.81. Median value was 1.96 and median 0.95 quartile was 2.75. This indicated that the epidemic was unabated and probably reflected the basic reproduction number of SARS-Cov-2 (table 1). The value of *R*_*i*_ at 21 days after the first case was registered in each state ranged from 1.09 to 1.89 (median 1.32 and 0.95 quartile median 1.5), indicating that the proliferation rate of the disease was diminishing, and the virus spread was slower (table 2). The EpiEstim package output includes an epidemic curve and a graph of the estimated *R*_*i*_ value, with 95% confidence intervals. The inspection of the *R*_*i*_ graphs could give some insight into the regional differences in COVID-19 infection spread (figures 1-3).

**Table 1:** Estimated instantaneous R at day 7 after first case of COVID-19 registered on each state of Brazil. The R values correlate well with the regions that would have higher case numbers and faster epidemic growth.

| State | Mean(R) | Std(R) | Quantile 0.05 | Quantile 0.95 |
| --- | --- | --- | --- | --- |
| AC | 1.76 | 0.37 | 1.31 | 2.28 |
| AL | 1.89 | 0.53 | 1.12 | 2.83 |
| AM | 2.81 | 0.61 | 1.92 | 3.94 |
| AP | 2.0 | 0.52 | 1.26 | 2.93 |
| BA | 1.66 | 0.4 | 1.07 | 2.37 |
| CE | 2.3 | 0.5 | 1.62 | 3.26 |
| DF | 1.96 | 0.5 | 1.23 | 2.86 |
| ES | 1.9 | 0.51 | 1.14 | 2.79 |
| GO | 1.94 | 0.37 | 1.37 | 2.59 |
| MA | 1.99 | 0.42 | 1.37 | 2.73 |
| MG | 2.01 | 0.52 | 1.23 | 2.93 |
| MS | 1.97 | 0.38 | 1.42 | 2.64 |
| MT | 2.15 | 0.49 | 1.44 | 3.05 |
| PA | 1.9 | 0.41 | 1.3 | 2.65 |
| PB | 2.06 | 0.51 | 1.32 | 3.01 |
| PE | 2.49 | 0.62 | 1.61 | 3.58 |
| PI | 1.64 | 0.31 | 1.16 | 2.17 |
| PR | 1.55 | 0.25 | 1.18 | 2.0 |
| RJ | 2.51 | 0.56 | 1.67 | 3.5 |
| RN | 1.88 | 0.51 | 1.1 | 2.75 |
| RO | 1.92 | 0.42 | 1.3 | 2.71 |
| RR | 1.92 | 0.38 | 1.35 | 2.64 |
| RS | 1.75 | 0.36 | 1.21 | 2.4 |
| SC | 2.0 | 0.35 | 1.47 | 2.63 |
| SE | 2.01 | 0.41 | 1.42 | 2.75 |
| SP | 1.9 | 0.47 | 1.2 | 2.75 |
| TO | 2.11 | 0.5 | 1.41 | 3.03 |

**Table 2:** Estimated instantaneous R at day 21 after first case of COVID-19 registered on each state of Brazil. It is clear that R values markedly reduced across all country, paralleling the institution of lockdown.

| State | Mean(R) | Std(R) | Quantile 0.05 | Quantile 0.95 |
| --- | --- | --- | --- | --- |
| AC | 1.09 | 0.07 | 0.97 | 1.21 |
| AL | 1.38 | 0.18 | 1.1 | 1.68 |
| AM | 1.32 | 0.09 | 1.16 | 1.46 |
| AP | 1.89 | 0.3 | 1.45 | 2.4 |
| BA | 1.46 | 0.15 | 1.22 | 1.71 |
| CE | 1.34 | 0.09 | 1.2 | 1.47 |
| DF | 1.33 | 0.11 | 1.16 | 1.5 |
| ES | 1.33 | 0.16 | 1.09 | 1.6 |
| GO | 1.18 | 0.08 | 1.04 | 1.32 |
| MA | 1.46 | 0.15 | 1.23 | 1.71 |
| MG | 1.31 | 0.12 | 1.12 | 1.51 |
| MS | 1.18 | 0.08 | 1.05 | 1.32 |
| MT | 1.33 | 0.11 | 1.14 | 1.52 |
| PA | 1.51 | 0.16 | 1.27 | 1.77 |
| PB | 1.21 | 0.11 | 1.03 | 1.39 |
| PE | 1.24 | 0.09 | 1.09 | 1.39 |
| PI | 1.28 | 0.12 | 1.08 | 1.49 |
| PR | 1.29 | 0.09 | 1.15 | 1.43 |
| RJ | 1.62 | 0.21 | 1.3 | 1.98 |
| RN | 1.42 | 0.15 | 1.18 | 1.67 |
| RO | 1.46 | 0.17 | 1.19 | 1.75 |
| RR | 1.25 | 0.09 | 1.11 | 1.4 |
| RS | 1.32 | 0.11 | 1.14 | 1.49 |
| SC | 1.18 | 0.07 | 1.06 | 1.29 |
| SE | 1.22 | 0.12 | 1.03 | 1.42 |
| SP | 1.66 | 0.24 | 1.3 | 2.08 |
| TO | 1.31 | 0.14 | 1.1 | 1.56 |

**Figure 1:**
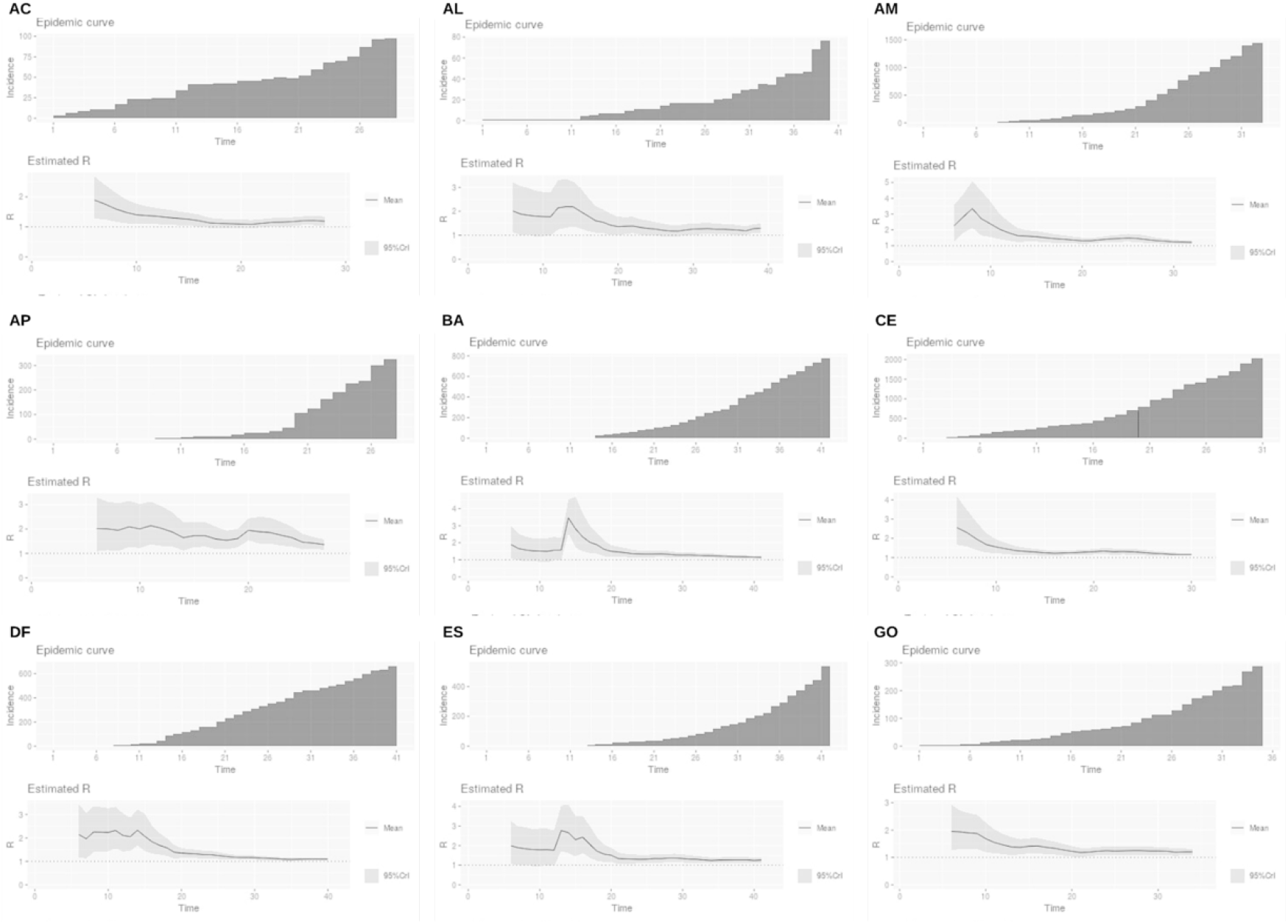
Epidemic curves and R_i_ estimate curves for some brazillian states: AC - Acre, AL - Alagoas, AM - Amazonas, AP - Amapá, BA - Bahia, CE - Ceará, DF - Distrito Federal, ES - Espírito Santo, GO - Goiás.

**Figure 2:**
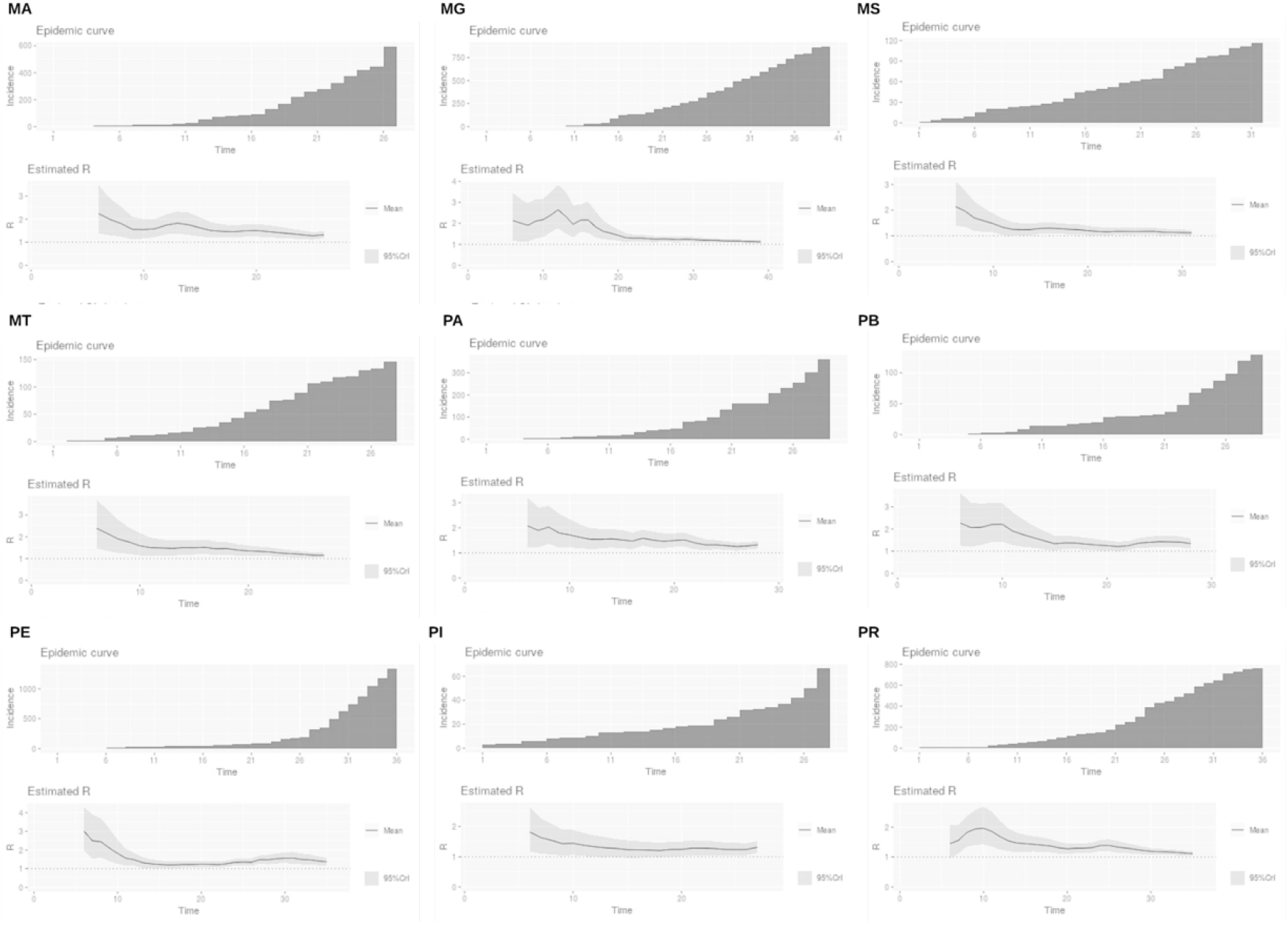
Epidemic curves and R_i_ estimate curves for some brazillian states: MA - Maranhão, MG - Minas Gerais, MS - Mato Grosso do Sul, MT - Mato Grosso, PA - Pará, PB - Paraíba, PE - Pernambuco, PI - Piaúi, PR - Paraná.

**Figure 3:**
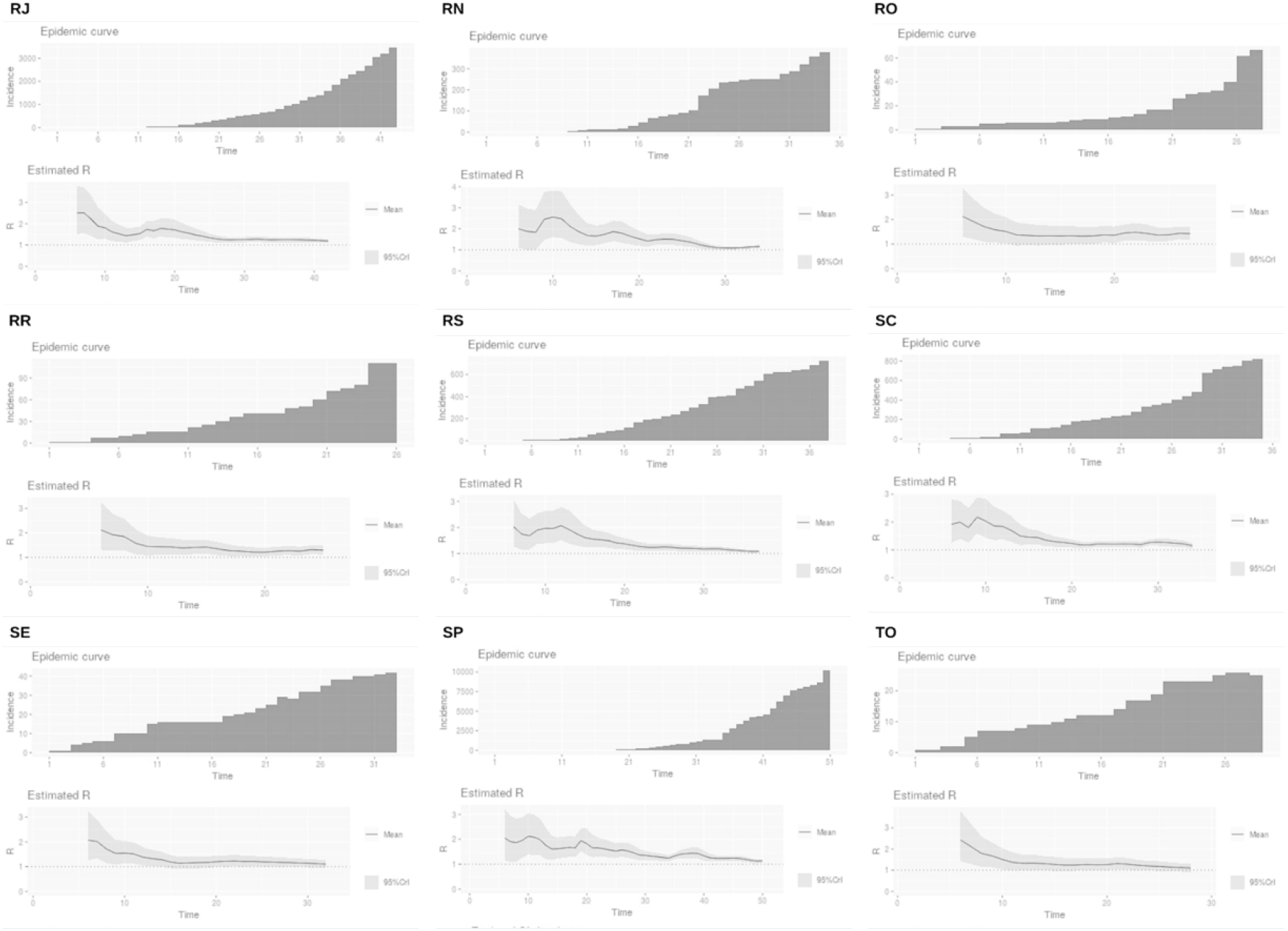
Epidemic curves and R_i_ estimate curves for some brazillian states: RJ - Rio de Janeiro, RN - Rio Grande do Norte, RO - Rondônia, RR - Roraima, RS - Rio Grande do Sul, SC - Santa Catarina, SE - Sergipe, SP - São Paulo, TO - Tocantins.

### Suplement

Links to data: - Ceará State - https://indicadores.integrasus.saude.ce.gov.br/api/casos-coronavirus/export-csv - São Paulo State - http://www.seade.gov.br/wp-content/uploads/2020/04/Dados-covid-19-estado.csv - Brazil - https://covid.saude.gov.br/

## Discussion

Our results showed an initial *R*_*i*_ compatible with the rapid epidemic growth rate in the beginning of the pandemic spread in Brazil, with most values *>* 2. However, 2 weeks later, the *R*_*i*_ showed a very impressive decline, reaching *<* 1.5 for the majority of brazillian states. This probably is a direct consequence of control measures instituted by local governments (social distancing, quarantine, lockdown). Furthermore, the highest *R*_*i*_ initial values correlated with the states that experienced high infection rates, like AM (Amazonas), RJ (Rio de Janeiro), and CE (Ceará).

In this study, we used incidence data to derive *R*_*i*_ and looked at brazillian states specific COVID-19 infection dynamics. In a model incorporating multiple variables, instantaneous R sensitively described real-time shifts of COVID-19 incidence, and varied accordingly with epidemic phase. Superspreading events were associated with *R*_*i*_ with high values, tipically much higher than 2, as well as rapid epidemic growth phases. In contrast, epidemic decline stage was characterized by *R*_*i*_ *<* 2 (Bandoy and Weimer, 2020).

The data on estimated *R*_*i*_ points to specific differences between different states. Some graphs exhibit spikes, like BA (Bahia) or ES (Espírito Santo) that may correlate with non compliance with control measures. Alternatively, local dynamics could play a role in determining *R*_*i*_ variation between different brazillian regions. Even in those regions that depicted variable behavior and *R*_*i*_ spikes, however, the trend was towards decreasing values and, therefore, less viral spread.

The basic reproduction number *R*_0_ is the expected number of infections caused by an individual in the absence of widespread immunity. Once widespread immunity is achieved, the effective reproduction number R will become lower than *R*_0_ and once R is less than 1, the population is said to have developed herd immunity and the epidemic declines. Immunity can only be obtained with certainty by vaccination, and a vaccine or effective treatment is not expected to be available soon. The best (and only) strategy right now is to rely on social distancing measures until a sustained epidemic suppression (R *<* 1) could be attained (Ferretti et al., 2020).

## Conclusion

Instantaneous R estimating showed to be a convenient way to investigate epidemic dynamics. The COVID-19 pandemic figures in Brazil indicate a trend towards the amellioration of the epidemic. However, this is heavily dependent upon regional governments decisions and politics. There is a high risk of new viral spread if the control measures are precociously eased. Authorities must take this into account and decide wisely upon social restrictions in the next few weeks, because they may be critical in this scenario.

## Data Availability

All data used in this research is public. Links were provided.

